# 3DeepVOG: An Open-Source Framework for Real-Time, Accurate 3D Gaze Tracking with Deep Learning

**DOI:** 10.1101/2025.11.01.25339065

**Authors:** Jingkang Zhao, Seyed-Ahmad Ahmadi, Julian Decker, Ken Möhwald, Peter zu Eulenburg, Andreas Zwergal, Virginia L. Flanagin, Max Wuehr

## Abstract

**Objective:** Eye movements are key biomarkers for diagnosing and monitoring neuro-otological, neuro-ophthalmological and neurodegenerative disorders. Video-oculography (VOG) systems enable detection of small, rapid eye movements and subtle oculomotor pathologies that may be missed during clinical exams. However, they rely on high-quality input for accurate tracking, struggle with torsional movements, and are often limited by high costs in broader clinical and research settings.

**Methods:** To overcome these limitations, we developed 3DeepVOG, a deep learning-based framework for three-dimensional monocular gaze tracking (horizontal, vertical, and torsional rotation) designed to operate robustly across varied imaging conditions, including low-light and noisy environments. The method includes automated framewise segmentation of the pupil and iris from video frames, followed by geometrically interpretable gaze estimation based on a two-sphere anatomical eyeball model incorporating corneal refraction correction. Torsion is tracked in real time using a novel mini-patch template matching approach. The system was trained on over 24,000 annotated samples obtained across multiple devices and clinical scenarios. Application was tested against a gold-standard VOG system in healthy controls.

**Results:** 3DeepVOG operates in real time (>300 fps) and achieves mean gaze errors of approximately 0.1° in all three motion dimensions. Derived oculomotor metrics – such as saccadic peak velocity, smooth pursuit gain, and optokinetic nystagmus slow-phase velocity – show good-to-excellent agreement with results from a clinical gold-standard system.

**Conclusions:** 3DeepVOG enables accurate, quantitative eye movement tracking across three dimensions under diverse conditions. As an open-source framework, it provides an accessible and scalable tool for advancing research and clinical assessment in neurological oculomotor disorders.

## 1. Introduction

Eye movements are critical neurophysiological signals and serve as biomarkers for diagnosing and monitoring neuro-otological disorders such as peripheral or central vestibular dysfunction, as well as central neurological system disorders such as multiple sclerosis and parkinsonian syndromes [1-4]. Governed by well-defined neural circuits, abnormalities often reflect specific lesions or dysfunctions in vestibular and oculomotor pathways. Structured clinical assessments evaluate mostly horizontal and vertical eye movements through tasks such as saccades, smooth pursuit, and optokinetic nystagmus (OKN), while torsional eye movements can be hardly evaluated without apparative testing [1, 5, 6]. Hypometric saccades and impaired pursuit are characteristic of neurodegenerative disorders such as progressive supranuclear palsy, while abnormal OKN – i.e., reduced slow-phase velocity or asymmetric responses – can indicate central vestibular or brainstem dysfunction. Torsional eye movements assessed for instance via the ocular tilt reaction, are crucial for diagnosing peripheral vestibular disorders and localizing lesions in the brainstem and cerebellum [7, 8].

Video-oculography (VOG) enables the detection of subtle or rapid eye movement abnormalities not evident on clinical examination, offering non-invasive, high-resolution tracking and the extraction of digital oculomotor biomarkers. Commercial VOG systems are widely used in clinical settings, but accuracy depends on infrared video quality, making them vulnerable to noise and poor lighting, especially during bedside or other less controlled environments [9-11]. Most commercial platforms focus on horizontal and vertical tracking and have limited sensitivity to torsional motion due to subtle iris textures and small rotational displacements. While scleral search coils allow precise torsion tracking, they are invasive and costly [12]. Additionally, the high cost and proprietary designs of current VOG systems limit broader clinical and research adoption.

Recent advances in computer vision and deep learning (DL) have enabled data-driven gaze tracking methods that are more robust to varying imaging conditions [10, 13, 14]. For instance, NVGaze, developed by NVIDIA, uses convolutional neural networks (CNNs) to estimate gaze directly from noisy raw VOG frames [13]. However, such end-to-end models often lack geometric interpretability – limiting clinical applicability where anatomical insight is crucial [15]. We previously introduced DeepVOG, among the first systems to combine DL with model-based gaze estimation for clinically interpretable gaze tracking [10, 16]. It uses a U-Net–like CNN for pupil segmentation and a calibration-free gaze estimator based on a simplified spherical eyeball model. However, its segmentation model (∼3,000 training frames) lacks robustness across conditions and devices, and the simplified spherical model, which ignores corneal refraction among other factors, limits anatomical accuracy [10].

To address these limitations, we present 3DeepVOG – an open-source system for robust 3D eye movement estimation (horizontal, vertical, and torsional) from VOG data. It features an enhanced segmentation network trained on a large, diverse dataset spanning multiple devices and clinical scenarios. 3DeepVOG integrates an anatomically accurate two-sphere eyeball model with corneal refraction correction and uses a GPU-accelerated, vectorized mini-patch template matching algorithm for real-time torsion tracking. In the following, we outline the methodology and evaluate real-time performance, segmentation accuracy, and 3D gaze estimation against clinical gold standards. By enabling accurate and interpretable oculomotor tracking under diverse conditions, 3DeepVOG provides a scalable, accessible tool for providing digital biomarkers of vestibular and neuro-ocular disorders.

## 2. Methods

### 2.1. System Pipeline

An overview of the 3DeepVOG pipeline, covering eye segmentation, and 3D gaze estimation are presented in Figure 1. Detailed mathematical and implementation-related information for each component of 3DeepVOG is provided in the Supplementary Appendix.

**Figure 1.**
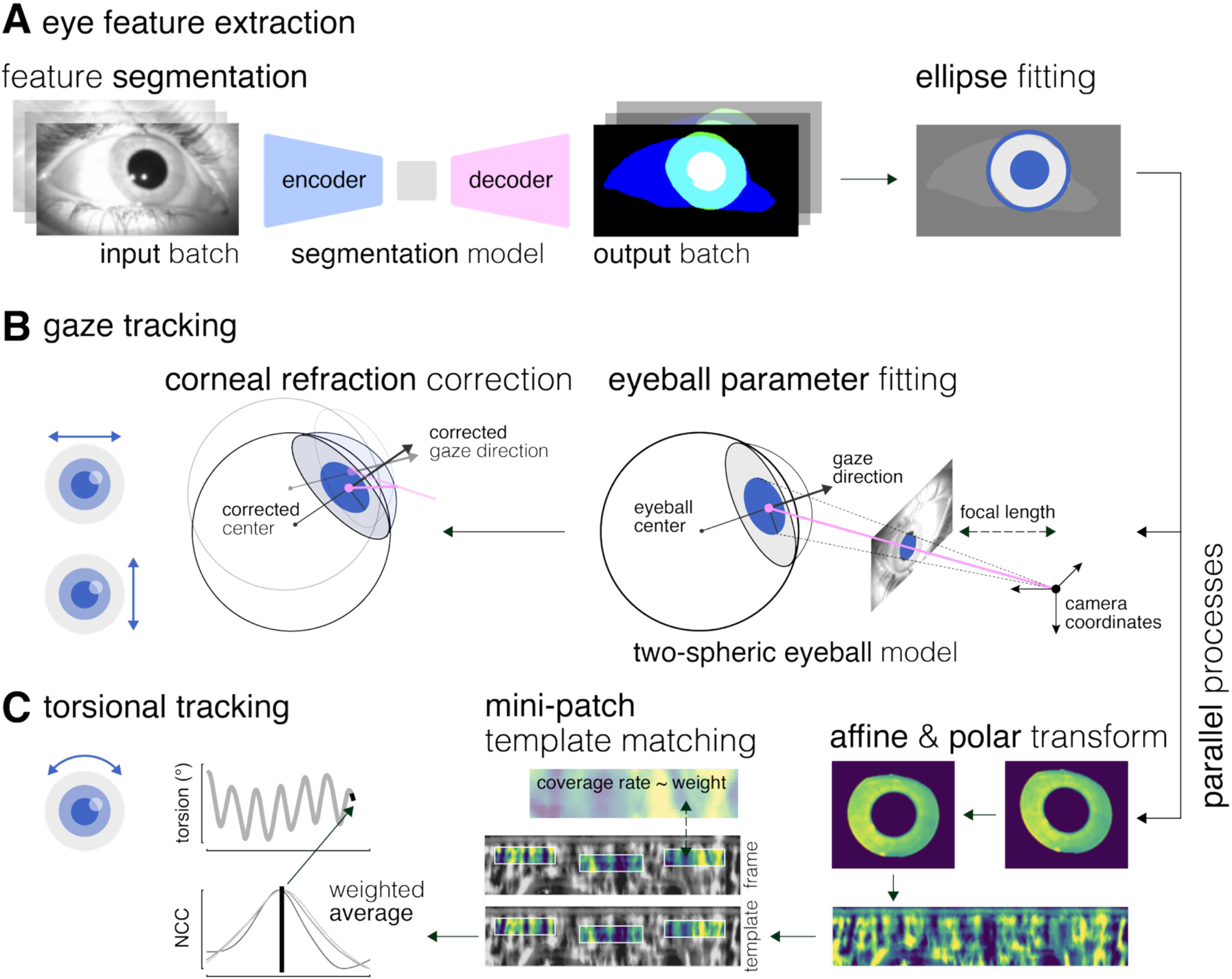
Overview of the 3DeepVOG System Pipeline. (A) Eye feature extraction: Input video frames are processed in mini-batches using a deep learning-based segmentation model, producing probability maps for the pupil, iris, and visible eye region. Post-processing yields fitted ellipses and segmented iris areas for downstream analysis. (B) Gaze tracking: Pupil ellipse parameters are used to fit a two-sphere anatomical eyeball model for estimating gaze vectors in the camera coordinate system followed by corneal refraction correction. (C) Torsion tracking: In a parallel process, the segmented iris region is geometrically corrected and unwrapped into polar coordinates. A mini-patch template matching algorithm is applied to track torsional eye movements by sampling local patches from the unwrapped iris map. For each patch the normalized cross-correlation (NCC) is used to measure similarity between the current patch position and its location in the template frame. Final torsion is computed by weighted average of mini-patch template matching results.

### 2.2. Eye Feature Extraction

#### 2.2.1. Segmentation Pipeline

Eye feature extraction begins with VOG frames as input (Fig. 1A). Each frame is resized and normalized to values between 0 to 1 (black to white). Frames with extreme brightness or darkness, identified by average pixel intensity outside predefined thresholds are excluded. The remaining valid frames are grouped into mini batches and passed through a DL segmentation model, generating probability maps for the pupil, iris, and visible eye regions. These maps are then resized back to the original frame resolution.

Postprocessing builds on an expanded DeepVOG pipeline [10]: Probability maps are binarized, and only the largest connected component is retained for each region. Ellipses are fitted to the pupil and iris masks to extract center coordinates, axis lengths, and rotation angles. The iris pattern map is computed by intersecting the fitted iris ellipse with the visible eye region and subtracting the pupil mask.

#### 2.2.2. Eye Segmentation Datasets

Training of the segmentation model was based on two datasets: the TEyeD dataset and a curated in-house dataset [10] all recorded from healthy volunteers under diverse conditions. TEyeD comprises VOG recordings across multiple devices and scenarios [17]. All TEyeD subsets were used except “NVGaze” which is no longer publicly available. Obvious annotation errors were excluded. Each subset was split approximately 60:20:20 into train, validation, and test sets, then concatenated and shuffled. Open-eye frames were downsampled to 70 Hz, resulting in 20,858 training, 7,213 validation, and 6,522 test frames.

The in-house dataset included manually labeled VOG frames from healthy adult participants recorded under diverse lighting environments (e.g., dark MRI, clinical, and natural light). Data were split 60:20:20 (3,523, 881, 1,102 frames) and balanced across TEyeD and in-house samples. Further recording details are provided in Supplementary Section 2.2.

#### 2.2.3. Model Comparison and Selection

We evaluated three lightweight encoder-decoder CNN architectures from MONAI [18], optimized for medical image segmentation: U-Net (1.63M parameters)[19], Attention U-Net (1.99M)[20], and SegResNet (1.58M)[21]. U-Net employs a 5-level encoder-decoder (16–256 channels) with skip connections and dropout (0.5); Attention U-Net adds attention gates; SegResNet uses residual blocks with batch normalization and dropout (0.2).

Segmentation performance was also compared with the prior DeepVOG model (24.7M; pupil-only)[10], and two VOG-specific baselines: RITnet (0.24M; no iris contour) [22] and EllSeg (2.6M; no visible eye region)[23] [23].

#### 2.2.4. Model Training and Evaluation

All candidate models were trained using a uniform preprocessing pipeline. The loss function combined Dice-Cross Entropy (λ=0.7) with a Hausdorff distance term (weight=0.03) to balance region and boundary accuracy. Optimization was performed using Adam (learning rate=0.004), with hyperparameters tuned via Optuna (100 trials on 10% of training data, each for 100 epochs) [24] .

Data augmentation (applied with 50% probability) included horizontal and vertical flips, affine transformations (±45° rotation, ±0.5 shear, ±15% translation), zoom (×0.9–2.0), gamma correction (γ=0.1–10), Gaussian noise (μ=0, σ=1), grayscale-to-RGB conversion, intensity scaling, and frame resizing.

Test performance was assessed on 3,843 VOG frames (including 1,511 with blinks) using multiple metrics: Dice coefficient, Hausdorff distance, precision, recall, and F1 score, computed for pupil, iris, and visible iris regions.

### 2.3. Blink Detection

Blinks were detected using a confidence score defined as the ratio between visible and total pupil area within the visible eye region. Frames with a score below 0.735 (determined via grid search) were classified as blinks. Detection performance was validated using annotated blink intervals from the TEyeD dataset (see Section 2.2.2).

### 2.4. Horizontal and Vertical Gaze Tracking

For gaze estimation, 3DeepVOG employs a previously proposed single-camera, glint-free 3D method based on a two-sphere anatomical eye model with corneal refraction correction (Fig. 1B) [16, 25]. This model, adapted from Le Grand [26], represents the eye as two intersecting spheres, the eyeball and cornea, and accounts for pupil shape distortions due to corneal refraction. Each frame yields a pupil ellipse of varying size, from which two possible 3D pupil circles with arbitrary radius can be inferred. These candidates are projected back to the 2D image to estimate the eyeball center. The correct circle is selected by ensuring the projected gaze direction (perpendicular to the pupil circle) intersects with the projected eyeball center. The eyeball center is then optimized across frames by minimizing the deviation between projected and observed pupil positions, while maintaining the anatomical distance between pupil and eyeball centers. Gaze direction is subsequently computed per frame based on the pupil’s position relative to the optimized eyeball center, with corneal refraction corrected using an empirical polynomial model. Eyeball parameters are estimated from frames collected during a gaze calibration task (see Section 2.6.1) and are held fixed for all subsequent gaze tracking.

### 2.5. Torsional Gaze Tracking

#### 2.5.1. Preprocessing

To estimate torsional eye movements (Fig. 1C), the iris pattern map from the segmentation pipeline undergoes an affine transformation based on ellipse parameters (center, size, rotation) to correct for viewing-angle distortions without requiring an explicit 3D eyeball model. This yields a normalized map where the pupil and iris form concentric circles. The annular region between the pupil and iris is mapped to horizontal lines in the unwrapped rectangular iris image, where the upper and lower boundaries correspond to the pupil and iris ellipses. The affine transformation smoothly interpolates between pupil and iris parameters. The final unwrapped image has fixed angular × radial resolution, and pixel intensities are mapped using bilinear interpolation.

To enhance pattern visibility, adaptive histogram equalization is applied. Two quality metrics are then computed: iris coverage (visibility ratio) and pattern clarity (horizontal gradient intensity). The frame with the highest combined score is selected as the template for torsion tracking.

#### 2.5.2. Mini-Patch Template Matching

After preprocessing, torsional eye movements are estimated by identifying angular shifts between sequential unwrapped iris patterns. To enhance performance and improve robustness to occlusions, illumination changes, and image distortion, we propose a mini-patch template matching algorithm as an alternative to full-frame methods [5, 27, 28].

Small rectangular patches are randomly selected from the enhanced unwrapped iris map in each video frame, constrained to the central iris region, to avoid edge effects. Normalized cross-correlation is then used to measure similarity between the current patch position and its location in the template frame by horizontally shifting it within a range corresponding to ±15 degrees of physiologically expected torsional movement and identifying the position with the highest correlation score [27].

To ensure reliable estimates, shift values from each patch are weighted by the visible iris ratio. Only patches exceeding a visibility threshold (0.5 in our study) are included, and weights are normalized to sum to one. The final torsion estimate per frame is computed as a weighted combination of patch shifts.

### 2.6. 3D Gaze Tracking Validation Procedures

The accuracy of 3DeepVOG’s horizontal and vertical gaze tracking was evaluated in 19 healthy participants (mean age: 32.4 ± 7.0 years; 8 female). Torsional gaze tracking was evaluated in a separate group of 10 healthy participants (mean age: 32.1 ± 8.7 years; 5 female). All validation recordings were performed using the EyeSeeCam system (EyeSeeTec GmbH, Germany; 120 Hz) a clinically validated VOG platform that is widely established in clinical practice and research as a gold-standard reference.

For initial eye model fitting, we employed a free-viewing calibration paradigm in which participants kept their heads stationary while freely exploring the visual periphery, producing highly elliptical pupil shapes. To validate of horizontal and vertical gaze tracking, participants underwent a standard neuro-ophthalmological assessment including saccades, smooth pursuit, and optokinetic nystagmus (OKN). For torsion tracking validation, torsional eye movements were elicited using sinusoidal (1 Hz, ±3 mA) galvanic vestibular stimulation (GVS) via a constant-current stimulator (Model DS5, Digitimer, UK). Ground-truth torsion was derived from scleral marker tracking analyzed using the EyeSeeCam system.

Validation was performed on two levels. First, the accuracy of 3D eye position estimates was quantified based on the median and interquartile range of absolute angular errors relative to the gold standard (EyeSeeCam for gaze; scleral markers for torsion). For horizontal and vertical gaze, estimates from the proposed two-sphere model with corneal refraction correction were compared to a baseline single-sphere model (as used in DeepVOG [10]). For torsion, estimates from the proposed mini-patch template matching method were compared to a baseline whole-image template matching approach. Prior to analysis, torsional signals were detrended (moving average) and bandpass filtered (fourth-order Butterworth, 0.2–2.0 Hz).

Second, the validity of clinical biomarkers derived from 3DeepVOG was assessed against the gold standard. For 2D gaze tracking, standard oculomotor biomarkers – saccadic peak velocity, smooth pursuit gain, and OKN slow phase velocity (SPV) – were extracted from the neuro-ophthalmological assessment [29, 30]. For torsion tracking, magnitude-squared coherence between the GVS stimulus and the torsional response was analyzed [31, 32]. Agreement with the gold standard was evaluated using Pearson’s correlation coefficient and the intraclass correlation coefficient for absolute agreement (ICC(3,1)), interpreted according to established guidelines: poor (<0.5), moderate (0.5–0.75), good (0.75–0.9), and excellent (>0.9) agreement [33].

### 2.7. Computing Environment

All components were implemented in Python 3.11 and optimized for real-time performance using GPU acceleration (NVIDIA GeForce RTX 4090) and multi-threaded CPU (AMD Ryzen 9 7950X3D, 16 cores). Model training was conducted on the Clinical Open Research Engine (CORE) at LMU Klinikum. Key libraries included PyTorch, MONAI, OpenCV, scikit-image, pye3d, and Kornia.

## 3. Results

### 3.1. Eye Region Segmentation and Blink Detection Performance

Segmentation performance was evaluated across several models using spatial and global metrics, while also considering model size to account for computational complexity and potential inference trade-offs (Table 1). Among all tested architectures, SegResNet (1.58 million parameters) consistently achieved the best performance across pupil, iris, and visible iris regions. It also enabled reliable blink detection without explicit blink annotations during training, yielding a precision of 0.995, recall of 0.904, and F1-score of 0.966.

**Table 1.**
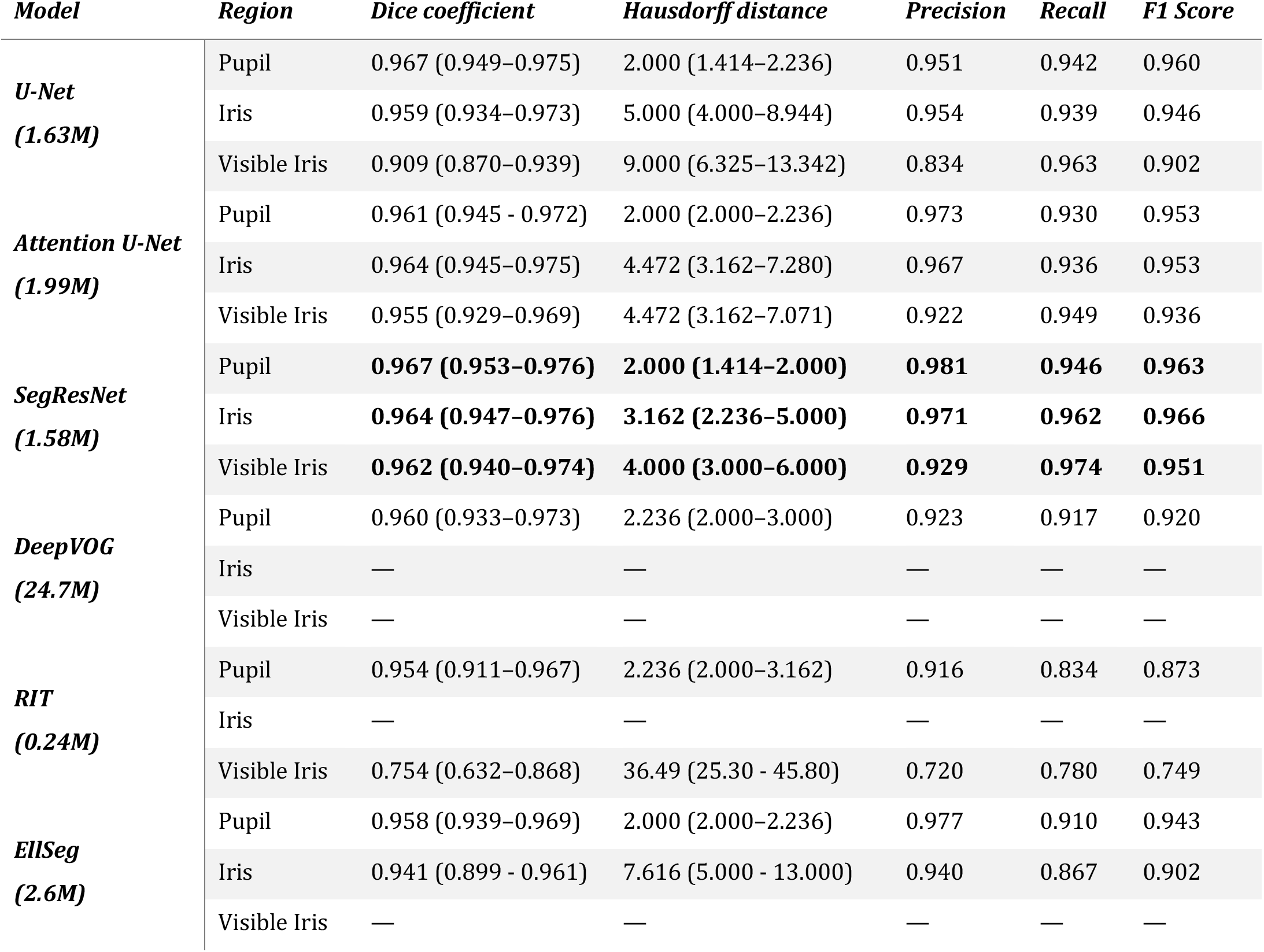
Eye region segmentation performance: Performance metrics for the evaluated segmentation networks (including number of network parameters) across different eye regions. Spatial differences between labeled and predicted regions are reported using Dice coefficient and Hausdorff distance (median and interquartile range), along with recall, precision, and F1-score. Best performance values are indicated in bold.

| *Model* | *Region* | *Dice coefficient* | *Hausdorff distance* | *Precision* | *Recall* | *F1 Score* |
| --- | --- | --- | --- | --- | --- | --- |
| **U-Net**<br>**(1.63M)** | Pupil | 0.967 (0.949–0.975) | 2.000 (1.414–2.236) | 0.951 | 0.942 | 0.960 |
|  | Iris | 0.959 (0.934–0.973) | 5.000 (4.000–8.944) | 0.954 | 0.939 | 0.946 |
|  | Visible Iris | 0.909 (0.870–0.939) | 9.000 (6.325–13.342) | 0.834 | 0.963 | 0.902 |
| **Attention U-Net**<br>**(1.99M)** | Pupil | 0.961 (0.945 - 0.972) | 2.000 (2.000–2.236) | 0.973 | 0.930 | 0.953 |
|  | Iris | 0.964 (0.945–0.975) | 4.472 (3.162–7.280) | 0.967 | 0.936 | 0.953 |
|  | Visible Iris | 0.955 (0.929–0.969) | 4.472 (3.162–7.071) | 0.922 | 0.949 | 0.936 |
| **SegResNet**<br>**(1.58M)** | Pupil | **0.967 (0.953–0.976)** | **2.000 (1.414–2.000)** | **0.981** | **0.946** | **0.963** |
|  | Iris | **0.964 (0.947–0.976)** | **3.162 (2.236–5.000)** | **0.971** | **0.962** | **0.966** |
|  | Visible Iris | **0.962 (0.940–0.974)** | **4.000 (3.000–6.000)** | **0.929** | **0.974** | **0.951** |
| **DeepVOG**<br>**(24.7M)** | Pupil | 0.960 (0.933–0.973) | 2.236 (2.000–3.000) | 0.923 | 0.917 | 0.920 |
|  | Iris | — | — | — | — | — |
|  | Visible Iris | — | — | — | — | — |
| **RIT**<br>**(0.24M)** | Pupil | 0.954 (0.911–0.967) | 2.236 (2.000–3.162) | 0.916 | 0.834 | 0.873 |
|  | Iris | — | — | — | — | — |
|  | Visible Iris | 0.754 (0.632–0.868) | 36.49 (25.30 - 45.80) | 0.720 | 0.780 | 0.749 |
| **EllSeg**<br>**(2.6M)** | Pupil | 0.958 (0.939–0.969) | 2.000 (2.000–2.236) | 0.977 | 0.910 | 0.943 |
|  | Iris | 0.941 (0.899 - 0.961) | 7.616 (5.000 - 13.000) | 0.940 | 0.867 | 0.902 |
|  | Visible Iris | — | — | — | — | — |

#### 3.2. 3D gaze tracking accuracy

3DeepVOG achieved median eye position errors of ∼0.1° across horizontal, vertical, and torsional planes (Table 2). For horizontal and vertical gaze, the proposed two-sphere anatomical model with corneal refraction correction significantly improved accuracy compared to the baseline single-sphere model, reducing errors by approximately 60–70% (Fig. 2A–D). For torsional position estimates, the mini-patch template matching method outperformed the baseline whole-image matching approach, achieving around 20% reduction in error (Fig. 2E–F).

**Table 2.**
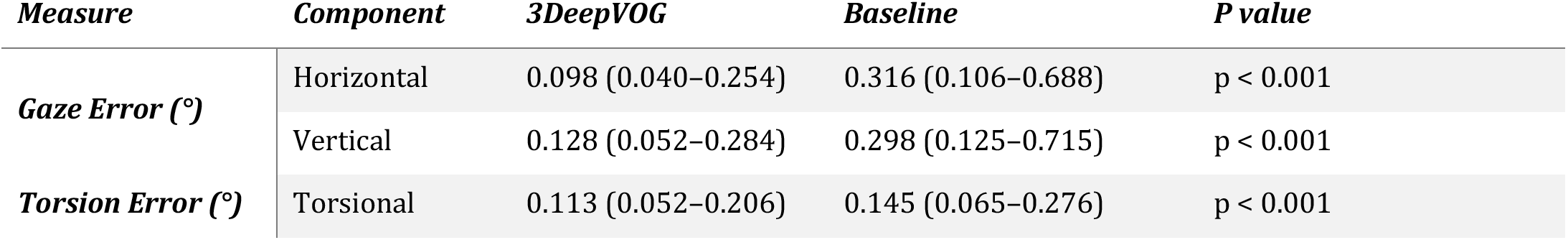
Accuracy of 3D eye position estimates. Absolute angular errors relative to the clinical gold standard are reported as median and interquartile range. Horizontal and vertical gaze errors are shown for the two-sphere eyeball model with corneal refraction correction used in 3DeepVOG, compared to a simplified one-sphere model without correction (baseline). Torsional position errors are shown for the mini-patch template matching algorithm implemented in 3DeepVOG, compared to a baseline full-template matching approach. Statistical differences between approaches are indicated in the last column.

| *Measure* | *Component* | *3DeepVOG* | *Baseline* | *P value* |
| --- | --- | --- | --- | --- |
| **Gaze Error (°)** | Horizontal | 0.098 (0.040–0.254) | 0.316 (0.106–0.688) | p < 0.001 |
|  | Vertical | 0.128 (0.052–0.284) | 0.298 (0.125–0.715) | p < 0.001 |
| **Torsion Error (°)** | Torsional | 0.113 (0.052–0.206) | 0.145 (0.065–0.276) | p < 0.001 |

**Figure 2.**
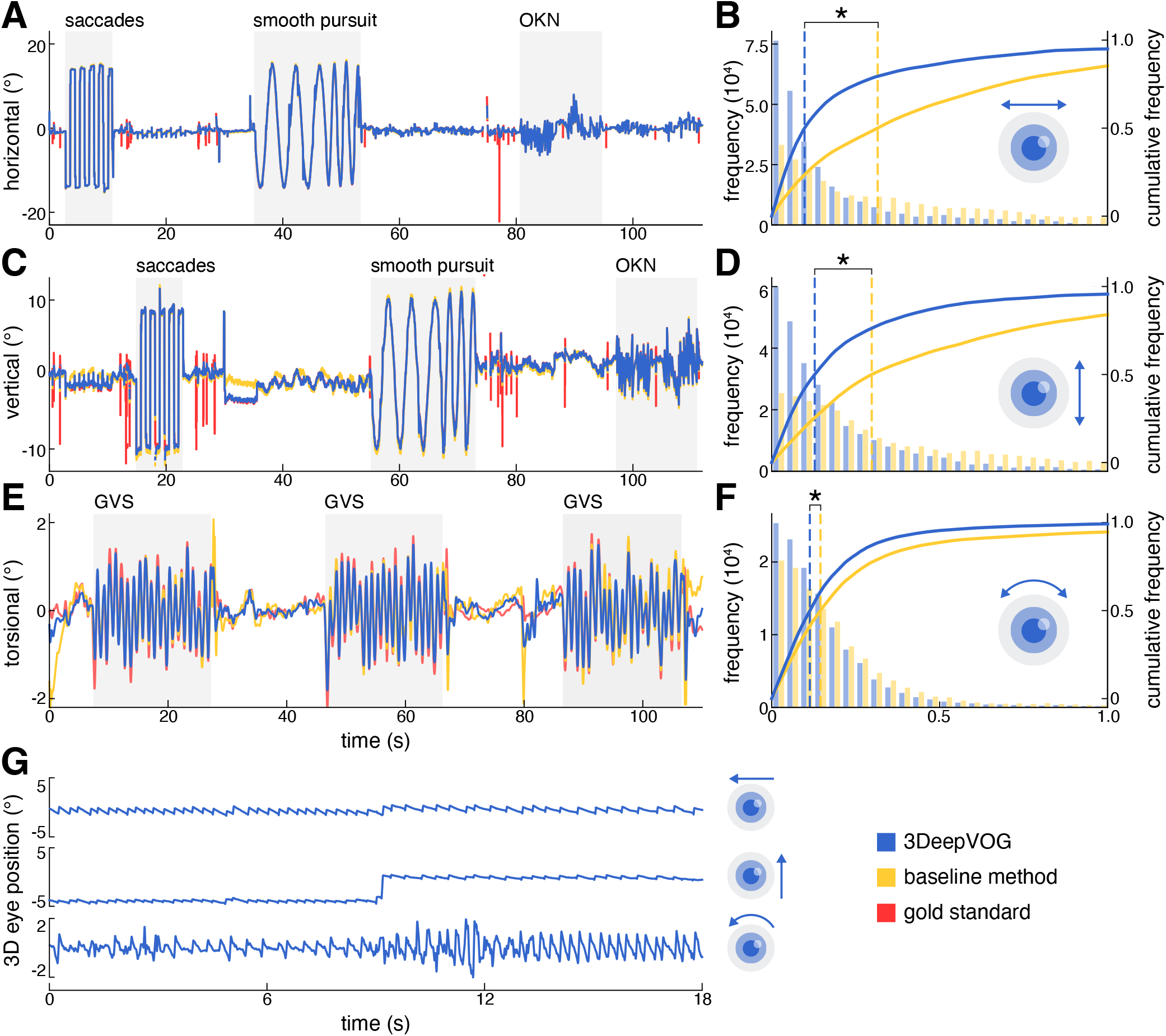
Accuracy of 3D eye position estimates. (A, C, E) Eye position traces derived from 3DeepVOG (blue) and a baseline method (yellow) are compared to the clinical gold standard (red). Horizontal and vertical eye movements were recorded during a standard oculomotor assessment including saccades, smooth pursuit, and optokinetic nystagmus (OKN). The baseline method for these components uses a simplified one-sphere eyeball model, whereas 3DeepVOG applies an anatomically accurate two-sphere model with corneal refraction correction. Torsional eye movements were recorded during repeated 1 Hz, ±3 mA galvanic vestibular stimulation (GVS). The baseline method applies full-template iris matching, while 3DeepVOG uses a mini-patch template matching approach. (B, D, F) Corresponding absolute angular error histograms and cumulative distribution functions (CDFs). Dashed lines indicate median errors for each method. Asterisks indicate statistically significant differences. (G) Exemplary 3D gaze tracking in a patient with acute unilateral vestibular failure, showing a right-beating and clockwise torsional nystagmus (from the patient’s perspective) with an up-beating component.

Across clinical oculomotor biomarkers (saccadic peak velocity, smooth pursuit gain, OKN SPV, and GVS– torsion coherence) 3DeepVOG showed good-to-excellent agreement with the clinical gold standard (EyeSeeCam), with ICC values ranging from 0.81 to 0.99 and relative magnitude errors (RMSE) below 8% (Fig. 3; Table 3).

**Table 3.** Validity of derived clinical oculomotor parameters. Comparison of oculomotor biomarkers derived from 3DeepVOG and EyeSeeCam. Values represent mean ± SD for each system, along with the absolute and relative root-mean-square error (RMSE), Pearson’s correlation coefficient (R, significant results in bold), and the intraclass correlation coefficient ICC (3,1). Abbreviations: OKN – optokinetic nystagmus; SPV – slow phase velocity; msCoherence – magnitude-squared coherence; GVS – galvanic vestibular stimulation.

| *Metric* | *EyeSeeCam*<br>(*mean ± SD*) | *3DeepVOG*<br>(*mean ± SD*) | *RMSE*<br>(*ABS*) | *RMSE*<br>(*REL %*) | *R* | *ICC (3,1)* |
| --- | --- | --- | --- | --- | --- | --- |
| ***Saccade Velocity***<br>***(°/s)*** | 412.499 ± 79.443 | 408.581 ± 81.104 | 22.604 | 5.48 | **0.961** | 0.961 |
| ***Smooth Pursuit Gain*** | 0.997 ± 0.083 | 0.985 ± 0.085 | 0.050 | 4.98 | **0.834** | 0.834 |
| ***OKN SPV (°/s)*** | 0.258 ± 8.112 | 0.251 ± 7.995 | 0.610 | 7.87 | **0.997** | 0.997 |
| ***msCoherence GVS-Torsion*** | 0.9245 ± 0.0824 | 0.9088 ± 0.0627 | 0.0457 | 4.94 | **0.845** | 0.818 |

**Figure 3.**
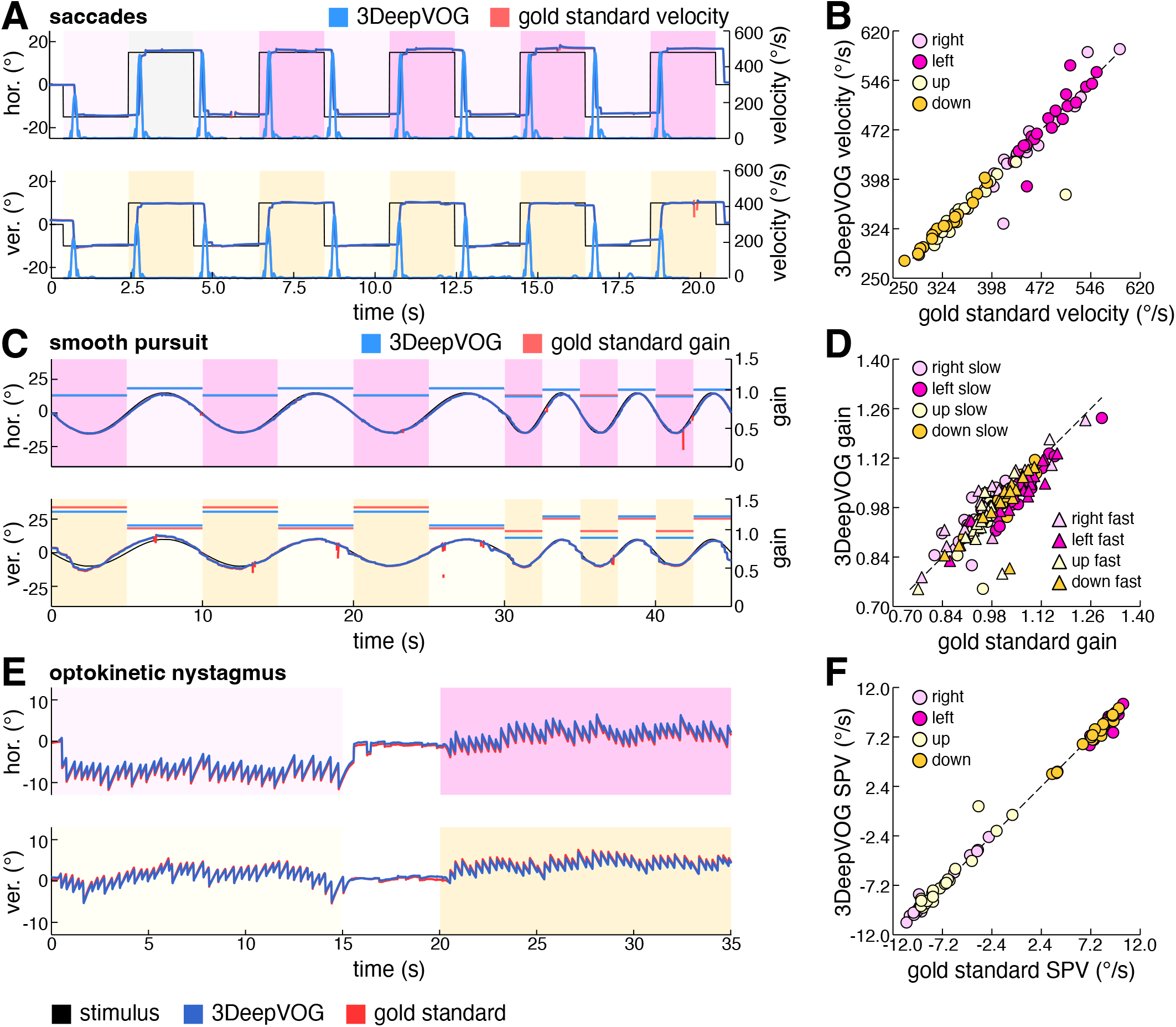
Validity of derived clinical oculomotor parameters. (A, C, E) Time-series plots of horizontal and vertical eye movements during saccades (A), smooth pursuit (C), and optokinetic nystagmus (OKN) (E), showing 3DeepVOG predictions (blue) alongside the clinical gold standard (red). Estimated biomarker values from 3DeepVOG (cyan) – saccade velocity (A) and pursuit gain (C) – are overlaid with corresponding gold standard values (magenta). (B, D, F) Correlation plots comparing 3DeepVOG-derived values with the clinical gold standard for saccade velocity (B), pursuit gain (D), and OKN slow-phase velocity (F). Markers indicate movement direction (left, right, up, down).

To demonstrate real-world applicability of 3DeepVOG built-in 3D gaze estimation pipeline, we analyzed data from a representative patient in their 70s with acute unilateral vestibular failure. Despite only moderate video quality, 3DeepVOG reliably captured the characteristic torsional eye movement patterns associated with this condition. The estimated three-dimensional nystagmus dynamics – comprising horizontal, vertical, and torsional components – were consistent with clinical expectations (Fig. 2G; Suppl. Video 1).

### 3.3. System Computational Performance

3DeepVOG achieved a preprocessing throughput of 300 fps, more than twice that of DeepVOG (120 fps), despite the additional computational demands of torsion tracking. This performance gain reflects the outlined architectural optimizations, multithreaded processing, and GPU acceleration.

## 4. Discussion

We introduced 3DeepVOG, an open-source framework for accurate, real-time binocular 3D gaze tracking. Building on our previous framework, DeepVOG – which at the time introduced the novel combination of DL-based eye region segmentation with anatomically grounded model fitting – 3DeepVOG implements several key improvements to the original framework. First, the segmentation network was retrained on a broader dataset comprising recordings from multiple VOG devices under diverse conditions. It now segments both the pupil and the iris, enabling accurate torsion estimation. We adopt SegResNet, a lightweight architecture that not only outperforms previous models in segmentation accuracy but also enables a reliable blink detection algorithm as part of the 3DeepVOG pipeline. Second, we introduce a refined two-sphere anatomical eye model with corneal refraction correction, improving horizontal and vertical gaze accuracy by up to 70% over DeepVOG and achieving ∼0.1° angular error. This level of precision lies within the sub-degree range typically required for clinical-grade and research eye-tracking systems, underscoring 3DeepVOG’s suitability for clinical and experimental applications [34]. Third, we introduce a mini-patch torsion-tracking method that improves both speed and accuracy over whole-image matching while retaining transparency and interpretability.

A key strength of 3DeepVOG is its adaptability to diverse VOG hardware without requiring specialized infrared or glint-based setups. Its reliable segmentation under natural lighting makes it well-suited for bedside, ambulatory, telemedical, and MRI-compatible applications. The framework’s accurate 3D eye-position estimation supports both clinical assessments and research requiring precise gaze localization. 3DeepVOG’s torsion tracking might be particularly relevant for research contexts where torsional eye movements act as proxies for vestibular responsiveness (e.g., binaural GVS) [35] and in neuro-otology and neuro-ophthalmology for assessing ocular tilt reactions or nystagmus characteristics [7, 8, 31] – though their clinical utility has so far been limited due to technical challenges in measurement. In this work, standard oculomotor biomarkers (saccadic peak velocity, smooth pursuit gain, and OKN SPV) derived from 3DeepVOG showed good-to-excellent agreement with a clinical gold standard, confirming the framework’s technical validity.

While 3DeepVOG offers robust and accurate 3D gaze tracking, several limitations remain. First all experiments were conducted in neurologically healthy participants, as this study represents a methodological validation focused on technical accuracy and robustness rather than diagnostic sensitivity or patient-level analysis. Follow-up evaluation studies are required to assess 3DeepVOG’s sensitivity and diagnostic applicability across diverse patient cohorts. Second, although DL is used for segmentation, many downstream steps rely on conventional methods, which may limit overall performance. Specifically, the current post-processing for estimating pupil and iris ellipses is CPU-based, creating computational bottlenecks due GPU-CPU data transfer. This could be mitigated by integrating these steps directly into the segmentation network, as previously demonstrated [23]. Similarly, anatomically informed gaze parameters could be learned from spatiotemporal input using transformer-based architectures [36]. Third, the torsion tracking algorithm works well with sharp iris images but fails under poor image quality or increased camera–eye distance [5, 27, 28]. A dedicated DL framework for torsion tracking could improve robustness [37]. Finally, 3DeepVOG is optimized for head-fixed eye-tracking systems used in research and clinics (e.g., EyeSeeCam, Pupil Labs Neon) and assumes known camera parameters. These assumptions currently limit its applicability in telemedical or remote monitoring scenarios, where the camera is positioned in front of the participant rather than mounted on the head, and the spatial relationship between the camera and the eye may change over time. Future work could integrate head-pose estimation based on facial landmark tracking to extend its use to remote monitoring via smartphones, tablets, or other consumer devices[38-40].

## Statement of Ethics

This work was performed in accordance with the Declaration of Helsinki. This human study was reviewed and approved by the Ethics Committee of the Medical Faculty, University of Munich, approval number 24-0108. All adult participants provided written informed consent prior to inclusion.

## Author Contributions

J.Z., S.A., P.E., A.Z., V.L.F., M.W. contributed to the conception and design of the study. J.Z., S.A., J.D., K.M., V.L.F., M.W. contributed to the methodology, acquisition of the data, J.Z. and M.W. wrote the draft of the manuscript. All authors contributed to the interpretation, writing, reviewing, and editing of the manuscript. All authors accepted responsibility for the integrity of the work and approved the final version of the manuscript.

## Data Availability Statement

3DeepVOG is freely available at https://github.com/DSGZ-MotionLab/3DeepVOG under the Apache-2.0 open-source license The repository includes pretrained models, example data, validation scripts, and detailed documentation with a step-by-step guide for installation and use. The complete validation data from this work can be obtained upon reasonable request from J.Z. Participants did not consent to the publication of their data in open repositories, in accordance with European data protection laws.

## Reference

1. Kassavetis, P., et al., Eye Movement Disorders in Movement Disorders. Movement Disorders Clinical Practice, 2022. 9(3): p. 284–295.

2. Sekar, A., M.T.N. Panouillères, and D. Kaski, Detecting Abnormal Eye Movements in Patients with Neurodegenerative Diseases - Current Insights. Eye and Brain, 2024. 16: p. 3–16.

3. Anderson, T.J. and M.R. MacAskill, Eye movements in patients with neurodegenerative disorders. Nature Reviews. Neurology, 2013. 9(2): p. 74–85.

4. Kheradmand, A., A.I. Colpak, and D.S. Zee, Eye movements in vestibular disorders. Handbook of Clinical Neurology, 2016. 137: p. 103–117.

5. Otero-Millan, J., et al., Knowing what the brain is seeing in three dimensions: A novel, noninvasive, sensitive, accurate, and low-noise technique for measuring ocular torsion. Journal of Vision, 2015. 15(14): p. 11.

6. Yu, Y., et al., Smooth Pursuit and Reflexive Saccade in Discriminating Multiple-System Atrophy With Predominant Parkinsonism From Parkinson’s Disease. Journal of Clinical Neurology, 2024. 20(2): p. 194–200.

7. Halmagyi, G.M., et al., Ocular tilt reaction: clinical sign of vestibular lesion. Acta Otolaryngol Suppl, 1991. 481: p. 47–50.

8. Otero-Millan, J., et al., The video ocular counter-roll (vOCR): a clinical test to detect loss of otolith-ocular function. Acta Otolaryngol, 2017. 137(6): p. 593–597.

9. Pleshkov, M., et al., Comparison of EOG and VOG obtained eye movements during horizontal head impulse testing. Frontiers in Neurology, 2022. 13.

10. Hoi, P.Y., C.Y. Cheung, and Z.T.H. Tse. DeepVOG: Open-source pupil segmentation and gaze estimation using deep learning. 2019.

11. Mantokoudis, G., et al., Impact of artifacts on VOR gain measures by video-oculography in the acute vestibular syndrome. J Vestib Res, 2016. 26(4): p. 375–385.

12. Houben, M.M., J. Goumans, and J. van der Steen, Recording three-dimensional eye movements: scleral search coils versus video oculography. Invest Ophthalmol Vis Sci, 2006. 47(1): p. 179–87.

13. Kim, J., et al. NVGaze: An Anatomically-Informed Dataset for Low-Latency, Near-Eye Gaze Estimation. in CHI ‘19: CHI Conference on Human Factors in Computing Systems. 2019. ACM.

14. Pathirana, P., et al., Eye gaze estimation: A survey on deep learning-based approaches. Expert Systems with Applications, 2022. 199: p. 116894.

15. Villanueva, A. and R. Cabeza, Models for Gaze Tracking Systems. EURASIP Journal on Image and Video Processing, 2007. 2007: p. 1–16.

16. Swirski, L. and N. Dodgson, A fully-automatic, temporal approach to single camera, glint-free 3D eye model fitting. 2013.

17. Fuhl, W., G. Kasneci, and E. Kasneci. TEyeD: Over 20 million real-world eye images with Pupil, Eyelid, and Iris 2D and 3D Segmentations, 2D and 3D Landmarks, 3D Eyeball, Gaze Vector, and Eye Movement Types. 2021.

18. Cardoso, M.J., et al., MONAI: An open-source framework for deep learning in healthcare. 2022, arXiv.

19. Ronneberger, O., P. Fischer, and T. Brox, U-Net: Convolutional Networks for Biomedical Image Segmentation. 2015, arXiv.

20. Oktay, O., et al., Attention U-Net: Learning Where to Look for the Pancreas. 2018, arXiv.

21. Myronenko, A., 3D MRI brain tumor segmentation using autoencoder regularization. 2018, arXiv.

22. Chaudhary, A.K., et al. RITnet: Real-time Semantic Segmentation of the Eye for Gaze Tracking. 2019.

23. Kothari, R.S., et al., EllSeg: An Ellipse Segmentation Framework for Robust Gaze Tracking. IEEE Transactions on Visualization and Computer Graphics, 2021. 27(5): p. 2757–2767.

24. Akiba, T., et al. Optuna: A Next-generation Hyperparameter Optimization Framework. 2019. Association for Computing Machinery.

25. Dierkes, K., M. Kassner, and A. Bulling. A fast approach to refraction-aware eye-model fitting and gaze prediction. in ETRA ‘19: 2019 Symposium on Eye Tracking Research and Applications. 2019. ACM.

26. Grand, Y.L., Light, Colour and Vision. 1968: Chapman & Hall.

27. Jin, N., et al., A Robust Method of Eye Torsion Measurement for Medical Applications. Information, 2020. 11(9): p. 408.

28. Ong, J.K. and T. Haslwanter, Measuring torsional eye movements by tracking stable iris features. J Neurosci Methods, 2010. 192(2): p. 261–7.

29. Leigh, R.J. and D.S. Zee, The Neurology of Eye Movements. 5 ed. 2015: Oxford University Press.

30. Kang, J.-J., et al., Recording and interpretation of ocular movements: saccades, smooth pursuit, and optokinetic nystagmus. Annals of Clinical Neurophysiology, 2023. 25(2): p. 55–65.

31. Jahn, K., et al., Inverse U-shaped curve for age dependency of torsional eye movement responses to galvanic vestibular stimulation. Brain, 2003. 126(Pt 7): p. 1579–89.

32. Mackenzie, S.W. and R.F. Reynolds, Ocular torsion responses to sinusoidal electrical vestibular stimulation. J Neurosci Methods, 2018. 294: p. 116–121.

33. Koo, T.K. and M.Y. Li, A Guideline of Selecting and Reporting Intraclass Correlation Coefficients for Reliability Research. J Chiropr Med, 2016. 15(2): p. 155–63.

34. Imai, T., et al., Comparing the accuracy of video-oculography and the scleral search coil system in human eye movement analysis. Auris Nasus Larynx, 2005. 32(1): p. 3–9.

35. Rühl, M., et al., In Vivo Localization of the Human Velocity Storage Mechanism and Its Core Cerebellar Networks by Means of Galvanic-Vestibular Afternystagmus and fMRI. The Cerebellum, 2022. 22(2): p. 194–205.

36. Popovic, N., et al., Model-aware 3D Eye Gaze from Weak and Few-shot Supervisions. 2023, arXiv.

37. Mukunda, K., et al., Deep Learning Detection of Subtle Torsional Eye Movements: Preliminary Results. 2024.

38. Parker, T.M., et al., Proof of Concept for an “eyePhone” App to Measure Video Head Impulses. Digital Biomarkers, 2020: p. 1–8.

39. Friedrich, M.U., et al., Smartphone video nystagmography using convolutional neural networks: ConVNG. J Neurol, 2023. 270(5): p. 2518–2530.

40. Barahim Bastani, P., et al., Self-Recording of Eye Movements in Amyotrophic Lateral Sclerosis Patients Using a Smartphone Eye-Tracking App. Digit Biomark, 2024. 8(1): p. 111–119.

