## Supplementally Material for "3DeepVOG: An Open-Source Framework for Real-Time, Accurate 3D Gaze Tracking with Deep Learning"

**Supplementally Document**

### 1. Introduction

This supplementary document provides a detailed description of the mathematical formulations and algorithms implemented in 3DeepVOG. For eye feature segmentation (Section 2), we elaborate on the frame validity criteria based on brightness, the method for generating iris pattern maps, and the definition of blink confidence for assessing blink states. The dataset preparation process and model hyper parameter setting are also explained in detail. For gaze tracking (Section 3), the core algorithm and hyperparameters of the corneal-refraction-aware two-sphere eyeball model follow Swirski’s and Dierkes’s methods [1, 2], with minor improvements. We describe the eyeball fitting algorithm and gaze estimation process, including implementation of corneal refraction correction. For torsion tracking (Section 4), we outline the preprocessing steps for converting iris pattern maps and the use of template matching with mini patches.

### 2. Eye Feature Segmentation

#### 2.1. Segmentation Pipeline

Eye feature extraction begins with frame input $\mathcal{M}_{i}$ from video-oculography. Each frame is resized to $W_{\mathcal{F}}\times H_{\mathcal{F}}$= 320×240 and normalized to ${\tilde{\mathcal{M}}}_{i}\in\left[ 0,1 \right]^{W_{\mathcal{F}}\times H_{\mathcal{F}}}$. Frame validity is determined based on the mean intensity $v_{i}$:

$$\begin{aligned} v_{i}= \left\{ \begin{aligned} 0, &\frac{1}{W_{\mathcal{F}}H_{\mathcal{F}}}\sum_{x,y} {\tilde{\mathcal{M}}}_{i}\left( x,y \right)<T_{min} \cup\frac{1}{WH}\sum_{x,y} {\tilde{\mathcal{M}}}_{i}\left( x,y \right)>T_{max} \\ 1, &else \end{aligned} \right. , i=1,2,\ldots,N_{f}\# \end{aligned}\left( 1 \right)$$

Here, $T_{m\mathrm{in}}=0.05$ and $T_{m\mathrm{ax}}=0.95$. Valid frames are grouped into mini-batches (32 frames) and input into the segmentation model $\mathcal{F}$ (SegResNet [3]) producing probability maps for the pupil, iris, and visible eye region. These maps are resized back to the original resolution.

In postprocessing, each output frame (e.g. in frame $i$) is binarized at a threshold of 0.5, denoised by extracting the largest connected components, and used to generate clean binary masks for the pupil ($\mathcal{O}_{i}^{\mathrm{pup}}$), iris ($\mathcal{O}_{i}^{\mathrm{iris}}$), and visible eye region ($\mathcal{O}_{i}^{\mathrm{eye}}$). Ellipse fitting is applied based on the least-square method, yielding ellipse parameters$\mathcal{p}_{i}^{2D}=\left( c_{x,i}^{\mathrm{pup}},c_{y,i}^{\mathrm{pup}}, a_{i}^{\mathrm{pup}}, b_{i}^{\mathrm{pup}}, \theta_{i}^{\mathrm{pup}} \right), \mathcal{i}_{i}^{2D}=\left( c_{x,i}^{\mathrm{iris}},c_{y,i}^{\mathrm{iris}}, a_{i}^{\mathrm{iris}}, b_{i}^{\mathrm{iris}}, \theta_{i}^{\mathrm{iris}} \right)$ where $\left( c_{x,i},c_{y,i} \right)$ are center coordinates, $a_{i}$ and $b_{i}$ are semi major, minor axes, and $\theta_{i}$ is the rotation angle. Then visible iris map $\mathcal{U}_{i}$ is extracted by subtracting the pupil mask from the intersection of the iris ellipse mask and the visible eye region:

$$\begin{aligned} \mathcal{U}_{i}=\mathcal{M}_{i}⨀\left( \neg\mathcal{O}_{i}^{pup}\cap\mathcal{O}_{i}^{iris}\cap\mathcal{O}_{i}^{eye} \right)\#\left( 2 \right) \end{aligned}$$

A blink confidence score $C_{i}$ is then computed:

$$\begin{aligned} C_{i}=\frac{\sum\mathcal{O}_{i}^{pup}}{\sum\left( \mathcal{O}_{i}^{pup}\cap\mathcal{O}_{i}^{eye} \right)}\#\left( 3 \right) \end{aligned}$$

Frames with $C_{i}\leq\theta^{\mathrm{blink}}$ are classified as blinks and excluded from further analysis. Here, $\theta^{\mathrm{blink}}$=0.735 determined empirically based on grid search.

#### 2.2. Eye Segmentation Datasets Details

The in-house segmentation data were obtained from healthy young adults in two cohorts: Dataset A (n = 35; 16 male, 19 female; 28.1 ± 4.0 years) recorded in a fully darkened MRI room, and Dataset B (n = 27; 8 male, 19 female; 25.5 ± 3.7 years) under normal illumination. MRI-compatible recordings were acquired using a NordicNeuroLab EyeTracking Camera (320 × 240 px, 60 Hz), supplemented with public datasets (Delhi[4] , LPW[5], MMU[6], PupilNet,[7] UBIRIS.v2[8]). All these data were collected from non-clinical populations based on their origional papers. The dataset appears to have been sampled non-sequentially, with missing index values indicating prior filtering. Dataset was randomly split into training, validation, and test sets in a 60:20:20 ratio (3,523, 881, 1102) randomly each those eight datasets to homogenize the sample size across train-validation-test datasets.

The TEyeD dataset integrates several healthy-subject subdatasets, we implemented a preprocessing pipeline to filter out error-prone video datasets based on criteria such as video file opening errors due to missing or corrupted files, runtime errors caused by memory issues or OpenCV failures, negative frame counts indicative of corrupted video files, and frame count mismatches between video and segmentation files. Additionally, datasets with incorrect annotations, such as inaccurate ellipse parameters, were discarded. To ensure balanced and structured data, valid videos were split training, validation, and test sets in a 60:20:20 ratio randomly, result in assigning each video to a single usage. The dataset was further refined through a down sampling strategy, sampling open-eye frames at 70Hz and blink frames at 5Hz for ensuring approximately 50% blink-prone images in the dataset. The video resources were converted into PNG images while annotation data was stored in three-channel NPY format, enabling efficient storage and faster access. Each dataset was cleaned to remove corrupted or mismatched annotations, then split 60:20:20 (train/val/test) ensuring every video was assigned to only one subset. Frames were downsampled (open-eye 70 Hz, blink 5 Hz) to balance blink and open-eye images.

### 3. Horizontal and Vertical Gaze Tracking

#### 3.1. Estimation Algorithm

During the eyeball fitting phase, for each frame, the 2D pupil ellipse parameters $\mathcal{p}_{i}^{2D}$ are used to estimate two possible unprojected pupil circles $\mathcal{p}_{i}^{\pm}\boldsymbol{=}\left( \boldsymbol{p}_{i}^{\pm},\boldsymbol{n}_{i}^{\pm}, r^{'} \right)\mathcal{=G}\left( \mathcal{p}_{i}^{2D}, f^{c}, r^{'} \right)$. Here $f^{c}$ is a known camera focal length, $r^{'}$ is an arbitrary predefined radiusand $\mathcal{G}$ is the reprojection function based on corneal geometry [2, 9]. By reprojecting these candidates, an initial 2D eyeball center is estimated, and an appropriate unprojected pupil circle $\mathcal{p}_{i}^{\boldsymbol{'}}=\left( \boldsymbol{p}_{i}^{\boldsymbol{'}},\boldsymbol{n}_{i}, r^{'} \right)$ is selected for each frame based on geometric consistency [2].

In the pye3D library, the 3D eyeball center $\boldsymbol{c}_{i}$ at the $i$-th frame is estimated by minimizing cumulative projection errors under anatomical constraints. This estimation uses three different time scales (short-term, long-term, and ultra-long-term) each producing a candidate center. The final $\boldsymbol{c}_{i}$ is computed from their interaction to ensure stable and accurate tracking over time [1, 10, 11]. Once $\boldsymbol{c}_{i}$ stabilizes (i.e., remains unchanged for over 80 frames), it is stored as the candidate fitted center $\boldsymbol{c'}$. The pupil center $\boldsymbol{p}_{i}$ is estimated such that $\left\| \boldsymbol{p}_{i}-\boldsymbol{c'} \right\|=d_{p}$ where $d_{p}$ is the anatomical pupil-to-eyeball distance.

To evaluate model fitting, two binary iris ellipse maps $\mathcal{E}_{i}^{\mathrm{iris}}$ and ${\tilde{\mathcal{E}}}_{i}^{\mathrm{iris}}$ are generated from the estimated 2D iris parameters $\mathcal{i}_{i}^{2D}$ and projected iris ring $\mathcal{i}_{i}^{\mathrm{Model}}$ of the eyeball model based on candidate fitted center $\boldsymbol{c'}$ and pupil center $\boldsymbol{p}_{i}$. Their overlap is measured by the Dice coefficient $\mathcal{d}_{i}$:

$$\begin{aligned} \mathcal{d}_{i}=\frac{2\left| \mathcal{E}_{i}^{iris}\cap{\tilde{\mathcal{E}}}_{i}^{iris} \right|}{\left| \mathcal{E}_{i}^{iris} \right|+\left| {\tilde{\mathcal{E}}}_{i}^{iris} \right|}\#\left( 4 \right) \end{aligned}$$

If $\mathcal{d}_{i}>\theta^{\mathrm{fit}}$, (empirically set $\theta^{\mathrm{fit}}$=0.95), the fit is accepted, and $c'$ is saved as the final 3D eyeball center $\mathbf{c}$ for subsequent gaze estimation.

During gaze estimation phase, the gaze vector for each frame is estimated as $\boldsymbol{n}_{i}=\frac{\boldsymbol{p}_{i}-\boldsymbol{c}}{\left\| \boldsymbol{p}_{i}-\boldsymbol{c} \right\|_{2}}$. Corneal refraction is corrected using empirical polynomial functions $\mathcal{R}_{\mathrm{eye}}$ and $\mathcal{R}_{\mathrm{gaze}}$ from pye3d library, yielding the corrected eyeball center $\tilde{\boldsymbol{c}}\boldsymbol{=}\mathcal{R}_{\mathrm{eye}}\left( \boldsymbol{c,}n_{\mathrm{ref}} \right)$**,** and gaze vector ${\tilde{\boldsymbol{n}}}_{i}\boldsymbol{=}\mathcal{R}_{\mathrm{gaze}}\left( \tilde{\boldsymbol{c}}\boldsymbol{,}\boldsymbol{n}_{i}, n_{\mathrm{ref}} \right)$**,** where $n_{\mathrm{ref}}$ is refractive index of aqueous humor.

#### 3.2. Postprocessing

Since the estimated gaze vectors ${\tilde{\boldsymbol{n}}}_{i}$ are expressed in the camera coordinate system, while the gaze coordinates from the clinical gold standard reference (EyeSeeCam) are provided in an unknown world frame, we optimized a linear transformation matrix $\boldsymbol{M\in}\mathbb{R}^{3\times3}$ to align them. The horizontal ($\phi_{i}$) and vertical ($\theta_{i}$) gaze components at frame i were computed as:

$$\begin{aligned} \left( \phi_{i}, \theta_{i} \right)\mathcal{=T}\left( {\tilde{\boldsymbol{n}}}_{i} \right)=\left( {tan}^{-1} \left( \frac{n_{z,i}}{n_{x,i}} \right)+\frac{\pi}{2}, {cos}^{-1} \left( \frac{n_{z,i}}{n_{y,i}} \right)-\frac{\pi}{2} \right)\#\left( 5 \right) \end{aligned}$$

where**,** ${\tilde{\boldsymbol{n}}}_{i}\boldsymbol{=}\left[ n_{x,i}, n_{y,i}, n_{z,i} \right]^{T}$**.** The transformation matrix $\boldsymbol{M}$ was optimized by minimizing the component-wise error between the predicted and ground truth angles:

$$\begin{aligned} M=\underset{\boldsymbol{M}^{\boldsymbol{'}}}{argmax} \sum_{i} \left\{ \left\| \mathcal{T}\left( \boldsymbol{M}^{\boldsymbol{'}}{\tilde{\boldsymbol{n}}}_{i} \right)_{\phi}\boldsymbol{-}\phi_{i}^{GT} \right\|+\left\| \mathcal{T}\left( \boldsymbol{M}^{\boldsymbol{'}}{\tilde{\boldsymbol{n}}}_{i} \right)_{\theta}\boldsymbol{-}\theta_{i}^{GT} \right\| \right\}\#\left( 6 \right) \end{aligned}$$

where $\phi_{i}^{\mathrm{GT}}, \theta_{i}^{\mathrm{GT}}$are the horizontal and vertical gold standard reference gaze coordinates.

### 4. Torsional Gaze Tracking

#### 4.1. Preprocessing

The original iris pattern map $\mathcal{U}_{i}$ is reversely mapped from an intermediate image ${\tilde{\mathcal{U}}}_{i}$ where the pupil, iris, and surrounding iris ring are normalized to appear circular. A linear transformation approximates the mapping back to the elliptical geometry. The inner and outer radii of the iris are computed from the fitted ellipse parameters: $r_{\mathrm{in}}=\frac{a_{i}^{\mathrm{pup}}+b_{i}^{\mathrm{pup}}}{2}$, $r_{\mathrm{out}}=\frac{a_{i}^{\mathrm{iris}}+b_{i}^{\mathrm{iris}}}{2}$. From ${\tilde{\mathcal{U}}}_{i}$, the iris ring is unwrapped into a polar map $\mathcal{V}_{i}\in\mathbb{R}^{W_{\theta}\times H_{r}}$ with predefined angular and radial resolutions: $W_{\theta}=3600$[pixels] and $H_{r}=50$[pixels]. Each pixel $\mathcal{V}_{i}\left( h, w \right)$ corresponds to a point ${\tilde{\mathcal{U}}}_{i}\left( \tilde{x}_{w,h},\tilde{y}_{w,h} \right)$ given by: $\tilde{x}_{w,h}=r_{h}\cos\left( \theta_{w} \right),\tilde{y}_{w,h}= r_{h}\sin\left( \theta_{w} \right)$ where $\theta_{w}=-\pi+w\delta_{\theta}\in\left[ -\pi,\pi\right], w=0,1,2,\ldots, W_{\theta}-1, \delta_{\theta}=\frac{2\pi}{W_{\theta}-1}$ and $r_{h}=r_{\mathrm{in}}+h\delta_{r}\in\left[ r_{\mathrm{in}},r_{\mathrm{out}} \right], h=0,1,2,\ldots H_{r}-1, \delta_{r}=\frac{r_{\mathrm{out}}-r_{\mathrm{in}}}{H_{r}-1}$.

At each radial step $h$, points on the circular ring: ${\tilde{\mathcal{U}}}_{i}\left( \tilde{x}_{w,h},\tilde{y}_{w,h} \right)$ $w=0,1,\ldots,W_{\theta}-1$ are mapped back to the corresponding elliptical ring in $\mathcal{U}_{i}\left( x_{w,h},y_{w,h} \right)$ by an affine transformation $\boldsymbol{A}_{h}^{\mathrm{affine}}\boldsymbol{\in}\mathbb{R}^{3\times3}$: $\left[ x_{w,h},y_{w,h}, 1 \right]^{T}=\boldsymbol{A}_{h}^{\mathrm{affine}}\left[ \tilde{x}_{w,h},\tilde{y}_{w,h}, 1 \right]^{T}$. The affine matrix is constructed as:

$$\begin{aligned} \boldsymbol{A}_{h}^{affine}=\boldsymbol{A}_{h}^{trans}\boldsymbol{A}_{h}^{rot}\boldsymbol{A}_{h}^{skew}\left( \boldsymbol{A}_{h}^{rot} \right)^{T}=\left[ \begin{matrix} 1 & 0 & c_{x,i}^{h} \\ 0 & 1 & c_{y,i}^{h} \\ 0 & 0 & 1 \end{matrix} \right]\left[ \begin{matrix} \cos\left( \theta_{i}^{h} \right) & -\sin\left( \theta_{i}^{h} \right) & 0 \\ \sin\left( \theta_{i}^{h} \right) & \cos\left( \theta_{i}^{h} \right) & 0 \\ 0 & 0 & 1 \end{matrix} \right]\left[ \begin{matrix} s_{x,i}^{h} & 0 & 0 \\ 0 & s_{y,i}^{h} & 0 \\ 0 & 0 & 1 \end{matrix} \right]\left[ \begin{matrix} \cos\left( -\theta_{i}^{h} \right) & -\sin\left( -\theta_{i}^{h} \right) & 0 \\ \sin\left( -\theta_{i}^{h} \right) & \cos\left( -\theta_{i}^{h} \right) & 0 \\ 0 & 0 & 1 \end{matrix} \right]\#\left( 7 \right) \end{aligned}$$

where $\boldsymbol{A}_{h}^{\mathrm{trans}}$ translates to the ellipse center, $\boldsymbol{A}_{h}^{\mathrm{rot}}$ rotates to align the ellipse axes, and $\boldsymbol{A}_{h}^{\mathrm{skew}}$ scales the circle into an ellipse. The scaling factors that define the ellipse’s skewness are computed as $s_{x,i}^{h}=\frac{2a_{i}^{h}}{a_{i}^{h}+b_{i}^{h}}, s_{y,i}^{h}=\frac{2b_{i}^{h}}{a_{i}^{h}+b_{i}^{h}}$. The transformation parameters $\gamma_{i}^{h}=\left( c_{x,i}^{h},c_{y,i}^{h},s_{x,i}^{h},s_{y,i}^{h},\theta_{i}^{h} \right)$ are linearly interpolated between pupil and iris values: $\gamma_{i}^{h}=\gamma_{i}^{\mathrm{pup}}+\left( \gamma_{i}^{\mathrm{iris}}-\gamma_{i}^{\mathrm{pup}} \right)\delta_{h}\in\left[ \gamma_{i}^{\mathrm{pup}},\gamma_{i}^{\mathrm{iris}} \right] ,$ where, $\delta_{h}=\frac{h}{H_{r}-1}, h=0,1,2,\ldots H_{r}-1$.

Bilinear interpolation is used to populate the unwrapped iris map $\mathcal{V}_{i}\left( w,h \right)$ from non-integer positions in $\mathcal{U}_{i}\left( x_{w,h},y_{w,h} \right)$**.** To optimize the contrast, adaptive histogram equalization is applied to $\mathcal{V}_{i}$ producing an intensity-enhanced image ${\tilde{\mathcal{V}}}_{i}\in\left[ 0,1 \right]^{W_{\theta}\times H_{r}}$. A visible rate $\varsigma_{i}$ and intensity pattern score $\varrho_{i}$ are computed for the updated template iris pattern map $\mathcal{T}$ as:

$$\begin{aligned} \varsigma_{i}=\frac{\sum_{w,h} \boldsymbol{1}_{{\tilde{\mathcal{V}}}_{i}\left( w,h \right)>0}}{W_{\theta}H_{r}}\#\left( 8 \right) \end{aligned}$$

$$\begin{aligned} \varrho_{i}=\sum_{h=0}^{H_{r}-1} \sum_{w=0}^{W_{\theta}-2} \left| {\tilde{\mathcal{V}}}_{i}\left( w+1, h \right)-{\tilde{\mathcal{V}}}_{i}\left( w, h \right) \right| \cdot\boldsymbol{1}_{{\tilde{\mathcal{V}}}_{i}\left( w, h \right)>0}\#\left( 9 \right) \end{aligned}$$

Here, $\varsigma_{i}$ measures iris visibility area and $\varrho_{i}$ captures horizontal pattern intensity. Frames maximizing both metrics are selected as template maps. The frame that maximizes both metrics is selected as the template iris pattern map $\mathcal{T}$.

#### 4.2. Mini-Patch Template Matching

First, P rectangular patch seeds $\mathcal{s}_{p}=\left( w_{p},h_{p} \right)$ where $p\in\left\{ 1,2,\ldots P \right\}$, are randomly sampled from the enhanced unwrapped iris map ${\tilde{\mathcal{V}}}_{i}$ of each frame $i$. Sampling is constrained within the valid range: $\left( w_{p},h_{p} \right)\in\left\{ \left\lceil W_{P}/2 \right\rceil,\left\lceil W_{P}/2 \right\rceil+1,\ldots W_{\theta}-W_{P} \right\}\times\left\{ \left\lceil H_{P}/2 \right\rceil,\left\lceil H_{P}/2 \right\rceil+1,\ldots,H_{r}-\left\lceil H_{P}/2 \right\rceil\right\}$. Each sampled location defines a patch region $\mathbb{R}^{W_{P}\times H_{P}}\subset\mathbb{R}^{W_{\theta}\times H_{r}}$ , and the corresponding pixel set for patch $p$ is $\Omega_{p}=\left\{ \left( w,h \right)|w\in\left[ w_{p}-\left\lceil W_{P}/2 \right\rceil,w_{p}+\left\lceil W_{P}/2 \right\rceil\right],h\in\left[ h_{p}-\left\lceil H_{P}/2 \right\rceil,h_{p}+\left\lceil H_{P}/2 \right\rceil\right] \right\}.$Then, normalized cross-correlation (NCC) is computed between the patch in ${\tilde{\mathcal{V}}}_{i}$ and a reference template $\mathcal{T}$ over shifts $\tau\in\left\{ -\epsilon,-\epsilon+1\ldots,\epsilon\right\}$ where $\epsilon=$ $\left\lceil15\delta_{\theta} \right\rceil$ accounts for a ±15° maximum detectable torsional shift. The shifted patch is defined as: $\Omega_{p}\left( \tau\right)=\left\{ \left( w,h \right)|w\in\left[ w_{p}-\left\lceil W_{P}/2 \right\rceil+\tau,w_{p}+\left\lceil W_{P}/2 \right\rceil+\tau\right],h\in\left[ h_{p}-\left\lceil H_{P}/2 \right\rceil,h_{p}+\left\lceil H_{P}/2 \right\rceil\right] \right\}\subset\Omega_{p}$,

The NCC response at shift $\tau$ is:

$$\begin{aligned} \gamma_{i,p}\left( \tau\right)=\frac{\sum_{\left( w,h \right)\in\Omega_{p}\left( \tau\right)} \left( {\tilde{\mathcal{V}}}_{i}\left( w,h \right)-\mu_{p,{\tilde{\mathcal{V}}}_{i}}\left( \tau\right) \right)\left( \mathcal{T}\left( w,h \right)-\mu_{p,\mathcal{T}}\left( \tau\right) \right)}{\sqrt{\sum_{\left( w,h \right)\in\Omega_{p}\left( \tau\right)} \left( {\tilde{\mathcal{V}}}_{i}\left( w,h \right)-\mu_{p,{\tilde{\mathcal{V}}}_{i}}\left( \tau\right) \right)^{2}}\sqrt{\sum_{\left( w,h \right)\in\Omega_{p}\left( \tau\right)} \left( \mathcal{T}\left( w,h \right)-\mu_{p,\mathcal{T}}\left( \tau\right) \right)^{2}}}\in\left[ 0,1 \right]\#\left( 10 \right) \end{aligned}$$

where the local means are: $\mu_{p,{\tilde{\mathcal{V}}}_{i}}\left( \tau\right)=\frac{\sum_{\left( w,h \right)\in\Omega_{p}\left( \tau\right)} {\tilde{\mathcal{V}}}_{i}\left( w,h \right)}{\left| \Omega_{p}\left( \tau\right) \right|}, \mu_{p\mathcal{,T}}\left( \tau\right)=\frac{\sum_{\left( w,h \right)\in\Omega_{p}\left( \tau\right)} \mathcal{T}\left( w,h \right)}{\left| \Omega_{p}\left( \tau\right) \right|}$.

Patch weight $\tilde{\varsigma}_{i,p}$ is computed as: $\tilde{\varsigma}_{i,p}=\frac{\varsigma_{i,p}}{\sum_{p=1}^{P} \varsigma_{i,p}}, \mathrm{where} \varsigma_{i,p}=\frac{\sum_{\left( w,h \right)\in\Omega_{p}} \boldsymbol{1}_{{\tilde{\mathcal{V}}}_{i}\left( w,h \right)>0}}{\left| \Omega_{p} \right|}$.

Only patches satisfying $\tilde{\varsigma}_{i,p}>\theta_{p}$ (a predefined threshold) are considered valid. The final torsional estimate $\psi_{i}$ for frame $i$ s computed as the weighted sum of the peak correlation shifts across valid patches:

$$\begin{aligned} \psi_{i}=\sum_{i} \tilde{\varsigma}_{i,p}\underset{\tau}{argmax} \gamma_{i,p}\left( \tau\right)\boldsymbol{1}_{\varsigma_{i,p}>\theta_{p}}\#\left( 11 \right) \end{aligned}$$

**Reference**

1.	Dierkes, K., M. Kassner, and A. Bulling. *A fast approach to refraction-aware eye-model fitting and gaze prediction*. in *ETRA '19: 2019 Symposium on Eye Tracking Research and Applications*. 2019. ACM.

2.	Swirski, L. and N. Dodgson, *A fully-automatic, temporal approach to single camera, glint-free 3D eye model ﬁtting.* 2013.

3.	Myronenko, A., *3D MRI brain tumor segmentation using autoencoder regularization*. 2018, arXiv.

4.	Kumar, A. and A. Passi, *Comparison and combination of iris matchers for reliable personal authentication.* Pattern Recognition, 2010. **43**(3): p. 1016-1026.

5.	Tonsen, M., et al., *Labelled Pupils in the Wild: A Dataset for Studying Pupil Detection in Unconstrained Environments.* arXiv, 2015.

6.	Multimedia, U., *MMU Iris Dataset.* 2019.

7.	Fuhl, W., et al., *PupilNet: Convolutional Neural Networks for Robust Pupil Detection.* arXiv, 2016.

8.	Proença, H., et al., *The UBIRIS.v2: A Database of Visible Wavelength Iris Images Captured On-the-Move and At-a-Distance.* IEEE Transactions on Pattern Analysis and Machine Intelligence, 2010. **32**(8): p. 1529-1535.

9.	Safaee-Rad, R., et al., *Three-dimensional location estimation of circular features for machine vision.* IEEE Transactions on Robotics and Automation, 1992. **8**(5): p. 624-640.

10.	*pye3d Pupil Detection*. [cited 2025 23/05]; Available from: <https://docs.pupil-labs.com/core/developer/pye3d/>.

11.	*pye3d eyeball model github*. [cited 2025 23/05]; Available from: <https://github.com/pupil-labs/pye3d-detector/blob/master/pye3d/eye_model/base.py>.
